# The Potential role of Particulate Matter in the Spreading of COVID-19 in Northern Italy: First Evidence-based Research Hypotheses

**DOI:** 10.1101/2020.04.11.20061713

**Authors:** Leonardo Setti, Fabrizio Passarini, Gianluigi De Gennaro, Pierluigi Barbieri, Maria Grazia Perrone, Andrea Piazzalunga, Massimo Borelli, Jolanda Palmisani, Alessia Di Gilio, Prisco Piscitelli, Alessandro Miani

## Abstract

**Background:** An epidemic model based only on respiratory droplets and close contact could not fully explain the regional differences in the spread of the recent severe acute respiratory syndrome COVID-19 in Italy, which was fast and dramatic only in Lombardy and Po Valley. On March 16^th^ 2020, we presented a Position Paper proposing a research hypothesis concerning the association between higher mortality rates due to COVID-19 observed in Northern Italy and the peaks of particulate matter concentrations, frequently exceeding the legal limit of 50 µg/m^3^ as PM_10_ daily average

**Methods:** To assess environmental factors related to the spread of the COVID-19 in Italy from February 24^th^ to March 13^th^ (the date when the lockdown has been imposed over Italy), official daily data relevant to ambient PM_10_ levels were collected from all Italian Provinces between February 9^th^ and February 29^th^, taking into account the average time (estimated in 17 days) elapsed between the initial infection and the recorded COVID positivity. In addition to the number of exceedances of PM_10_ daily limit value, we considered also population data and daily travelling information per each Province.

**Results:** PM_10_ daily limit value exceedances appear to be a significant predictor (p < .001) of infection in univariate analyses. Less polluted Provinces had a median of 0.03 infection cases over 1000 residents, while most polluted Provinces had a median of 0.26 cases over 1000 residents. Thirty-nine out of 41 Northern Italian Provinces resulted in the category with highest PM_10_ levels, while 62 out of 66 Southern Provinces presented low PM_10_ concentrations (p< 0.001). In Milan, the average growth rate before the lockdown was significantly higher than Rome (0.34 vs. 0.27 per day, with a doubling time of 2.0 days vs. 2.6), suggesting a basic reproductive number R_0_>6.0, comparable with the highest values estimated for China.

## Introduction

Severe acute respiratory syndrome known as COVID-19 disease (due to SARS-CoV-2 virus), is recognized to spread via respiratory droplets and close contacts [1]. However, this unique transmission model does not seem to explain properly the different spread observed in Italy from February 24th, 2020 to March 13rd, 2020. The huge virulence of COVID19 in the Po Valley is not comparable to the milder contagiousness observed in the central-southern regions. Demographic factors related to the ageing of the population and the possibility of infection without clinical symptoms for a quite long time - associated with the high rate of asymptomatic people that characterize COVID-19, estimated in 50-75% of infections - may only partially explain the fast spreading of the virus in Lombardy and Northern Italy [2,3]. Cai et al (2020) reported different incubation periods in patient(s) infected in Wuhan [4], but an epidemic model based only on respiratory droplets and close contact could not fully explain the regional differences in the spreading of the recent severe acute respiratory syndrome COVID-19 in Italy, which was fast and dramatic only in Lombardy and Po Valley.

At the same time, a number of studies have shown that airborne transmission route could spread viruses even further the close contact with infected people [5-19]. Paules et al. (2020) highlighted that - besides close distance contacts - airborne transmission of SARS-CoV can also occur [5]. It has also been reported how for some pathogens the airborne transport can reach long distances [6-8]. Reche et al. (2018) described the aerosolization of soil-dust and organic aggregates in sea spray that facilitates the long-range transport of bacteria, and likely of viruses free in the atmosphere. In particular, virus deposition rates were positively correlated with organic aerosol <0.7 µm, implying that viruses could have longer persistence times in the atmosphere and, consequently, will be dispersed further [9]. Moreover Qin et al. (2020) analyzed the microbiome of the airborne particulate matter (PM_2.5_ and PM_10_) in Beijing over a period of 6 months in 2012 and 2013, putting in evidence a variability of the composition that depended on the months [10]. Temporal distribution of the relative abundance of the microbiome on the particulate matter (PM) showed the highest presence of viruses in January and February, just in coincidence with most severe PM pollution. Chen. et al (2017) demonstrated the relationship between short-term exposure PM_2.5_ concentration and measles incidence in 21 cities in China [11]. Their meta-analyses showed that the nationwide measles incidence was significantly associated with an increase of 10 µg/m^3^ in PM_2.5_ levels.

Other recent studies have also reported associations between PM and infectious diseases (e.g., influenza, hemorrhagic fever with renal syndrome): inhalation could bring PM deep into the lung and virus attached to particles may invade the lower part of respiratory tract directly, thus enhancing the induction of infections, as demonstrated by Sedlmaier et al (2009) [12]. Zhao et al. (2018) showed that the majority of the positive cases of highly pathogenic avian influenza (HPAI) H5N2 in Iowa (USA) in 2015 might have received airborne virus, carried by fine PM, from infected farms both within the same State and from neighboring States [13]. The condensation and stabilization of the bioaerosol, generating aggregates with atmospheric particles from primary (i.e. dust) and secondary particulate, has been indicated as mechanisms able to transport airborne bacteria and viruses to distant regions, even by the inter-continent-transported dust: Ma et al. (2017) observed a positive correlation of the measles incidence with PM_10_ in western China during the period 1986-2005 [14]; Ferrari et al. (2008) showed measles outbreaks occurring in dry seasons and disappearing at the onset of rainy seasons in Niger [15]; Brown et al (1935) found that the most severe measles epidemic in the United States occurred in Kansas in 1935 during the Dust Bowl period [16].

Coming to recent specific studies, laboratory experiments of Van Doremalen et al. (2020) indicated that airborne and fomite transmission of SARS-Cov-2 is plausible, since the virus can remain viable and infectious in aerosol for hours [17]. Field measurement by Liu et al. (2020) showed evidence of coronavirus RNA in air sampled in Wuhan Hospitals and even in ambient air in close proximity during COVID-19 outbreak, pointing at the airborne route as a possible important pathway for contamination, that should have a further confirmation [18]. Santarpia et. al. reported the presence of airborne SARS-Cov-2 in air sampled at the Nebraska University Hospital [19], while - at the opposite - some negative evidence of virus presence in air reported by Ong et al. (2020) come from explicitly poor sampling scheme [20]. A research carried out by the Harvard School of Public Health seems to confirm an association between increases in particulate matter concentration and mortality rates due to COVID-19 [21]. On March 16^th^ 2020, we have released an official Position Paper highlighting that there are enough evidence to consider airborne route as a possible additional factor for interpreting the anomalous COVID-19 outbreaks notified in the Northern Italy, known to be one of the European areas characterized by highest PM concentration [22,23]. Data that led to the publication of the Position Paper are presented in this article, and are expected to trigger the interest of the research community at working on this topic.

## Methods

We have analyzed daily data relevant to ambient PM_10_ levels, urban conditions and virus incidence from all Italian Provinces, in order to reliably determine the association between PM pollution level and the initial spread of COVID-19. PM_10_ daily concentration levels were collected by the official air quality monitoring stations of the Regional Environmental Protection Agencies, ARPA), publicly available on their websites. The number of PM_10_ daily limit value exceedances (50 μg/m^3^) detected in the different Provinces, divided by the total number of PM_10_ monitoring stations for each selected Province was taken into account. Population data, population density and number of commuters related to each Italian Province were collected from ISTAT database for the 110 Provinces [3]. The number of COVID-19 infected people for each Province from February 24^th^ to March 13^th^ (the date when the lockdown was decided) was that reported on the official Government website, updated with daily frequency [24]. PM_10_ exceedances were collected between February 9^th^ and February 29^th^, taking into account the *lag period*, which is the average time elapsed between the initial infection and the diagnosis. To investigate how high PM_10_ concentrations (above the daily limit value) might relate to infection diffusion, we performed an exploratory analysis considering the recursive binary partitioning tree approach, as implemented into the party package [25] of R [26].

Besides PM_10_ daily limit value exceedances we considered several further covariates related to the different Provinces: population absolute frequencies; population densities (n° inhabitants/km^2^); the absolute frequencies of people daily travelling as estimated by the Italian National Institute of Statistics [3], and its proportion with respect to the overall Province population. As response variable we considered the infection rate of the disease, expressed as a proportion obtained binding together into a single two-dimensional vector both the number of COVID-19 cases and the rest of the Province population. We have performed statistical inferences analyses on Milan and Rome data, in order to observe the potential association between PM levels and COVID-19 spreading in big cities located in different geographic areas and with remarkable differences in PM_10_ exceedances, presenting at the same time quite similar urbanization, life style, population, ageing index, and number of commuters.

## Results

The spatial distribution of ambient PM_10_ exceedances between Italian cities was geographically heterogeneous and it is presented in Fig. 1a. The highest numbers of exceedances were generally located in Northern Italian Regions, while zones with a lower contagion were sited in Central and Southern Regions.

**Figure 1.**
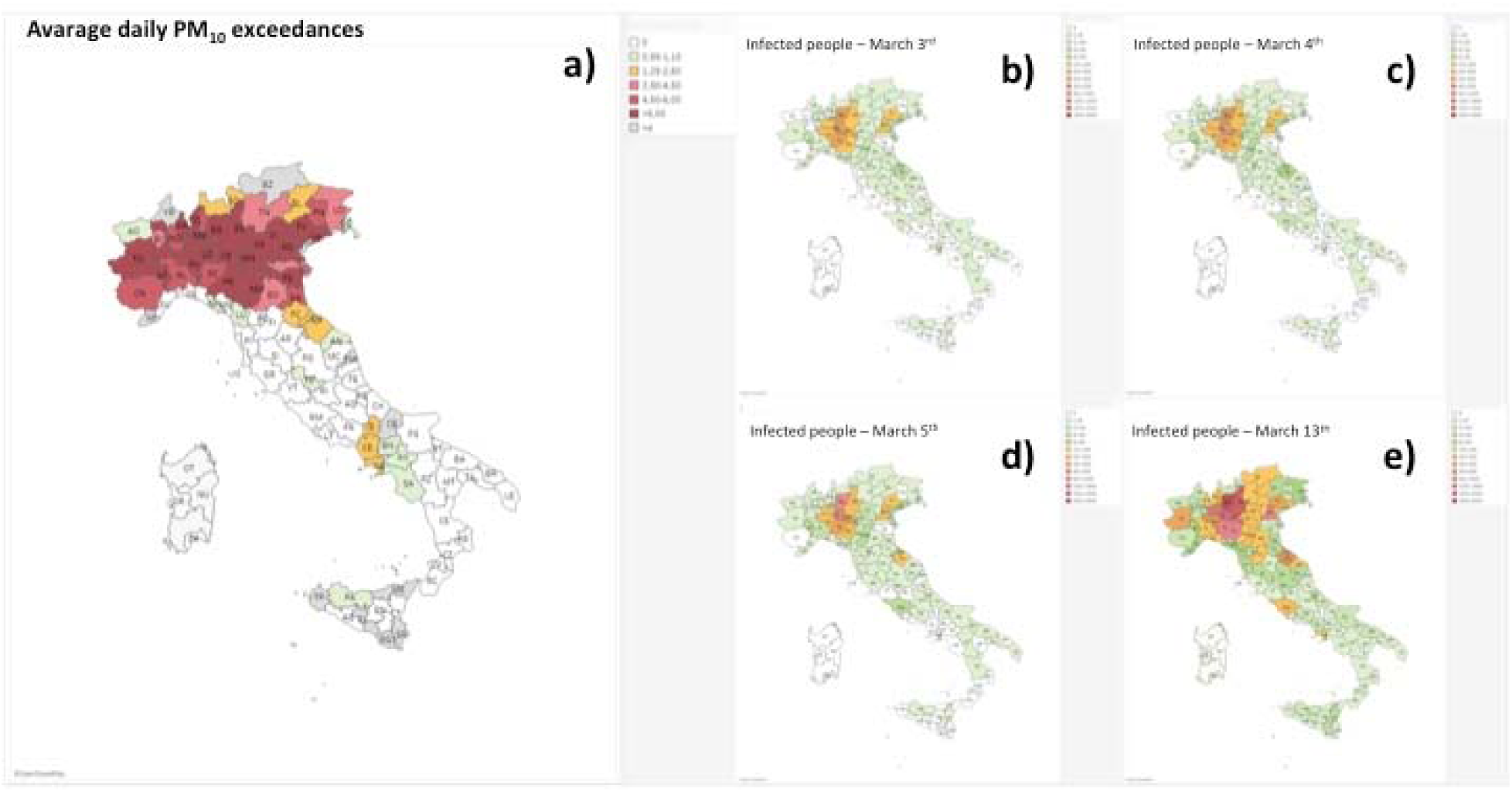
**(a)** Average daily PM_10_ exceedances vs. number of monitoring stations in different Italian Provinces from February 9^th^ to 29^th^ 2020; **(b-e)** Spreading of COVID-19 infected people during the period March 3^th^ – 13^th^ 2020

The maps in Fig 1 illustrate the mean of PM_10_ exceedances on the number of PM_10_ stations in all Italian Provinces in the period February 9^th^-29^th^ 2020 (Fig. 1a), compared with the total COVID-19 infection per Province observed in the period March 4^th^-13^th^ (Fig. 1b-e). Overall, there were 17,660 infected people during the time lapse of the study. The highest incidences of COVID-19 occurred in cities located in Northern Italy, and in particular in Lombardy Region, including its capital Milan. The lowest incidences of COVID-19 were observed in Southern Italy, as in Lazio Region, which includes Rome.

If continuing the observations beyond the date of the shutdown (March 13^th^), it was possible – by analyzing the trend of new daily COVID-19 infections - to observe a first reduction of the spreading rate of contagion around March 22^nd^ (reflecting the school closure ordered on March 5^th^) and a second one around March 28^th^ (reflecting the lockdown ordered on March 11^th^-13^th^) (Figure 2).

**Figure 2.**
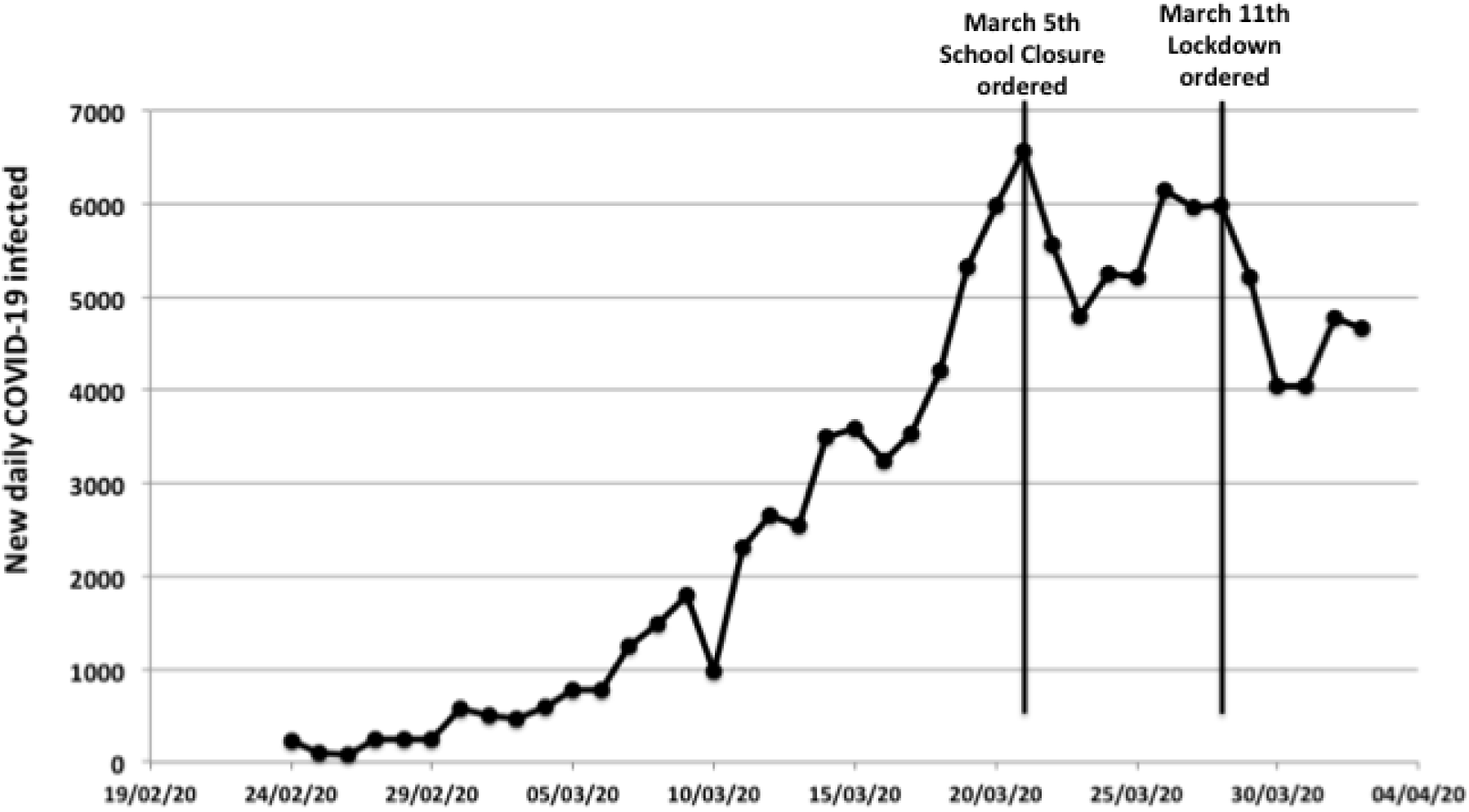
New daily COVID-19 infections in Italy from February 24^th^ to April 4^th^ 2020

On the basis, the *lag period* can be estimated in 17 days. In the univariate analysis, the PM_10_ daily limit value exceedances appear to be a significant predictor (p < .001) of infection (Fig. 3b) with a 1.29 cut-off value. The cut-off divides the Provinces into two classes, respectively with higher (n = 43) and lower (n = 67) PM_10_ concentrations. The boxplots depict the log-transformed infection rate of the disease: the less polluted Provinces had a median 0.03 infection case over 1000 residents (first – third quartile 0.01 – 0.09, range 0.00 – 0.56), while most polluted Provinces had a median 0.26 infection cases over 1000 Province residents (first – third quartile 0.14 – 0.51, range 0.00 – 4.92).

**Figure 3.**
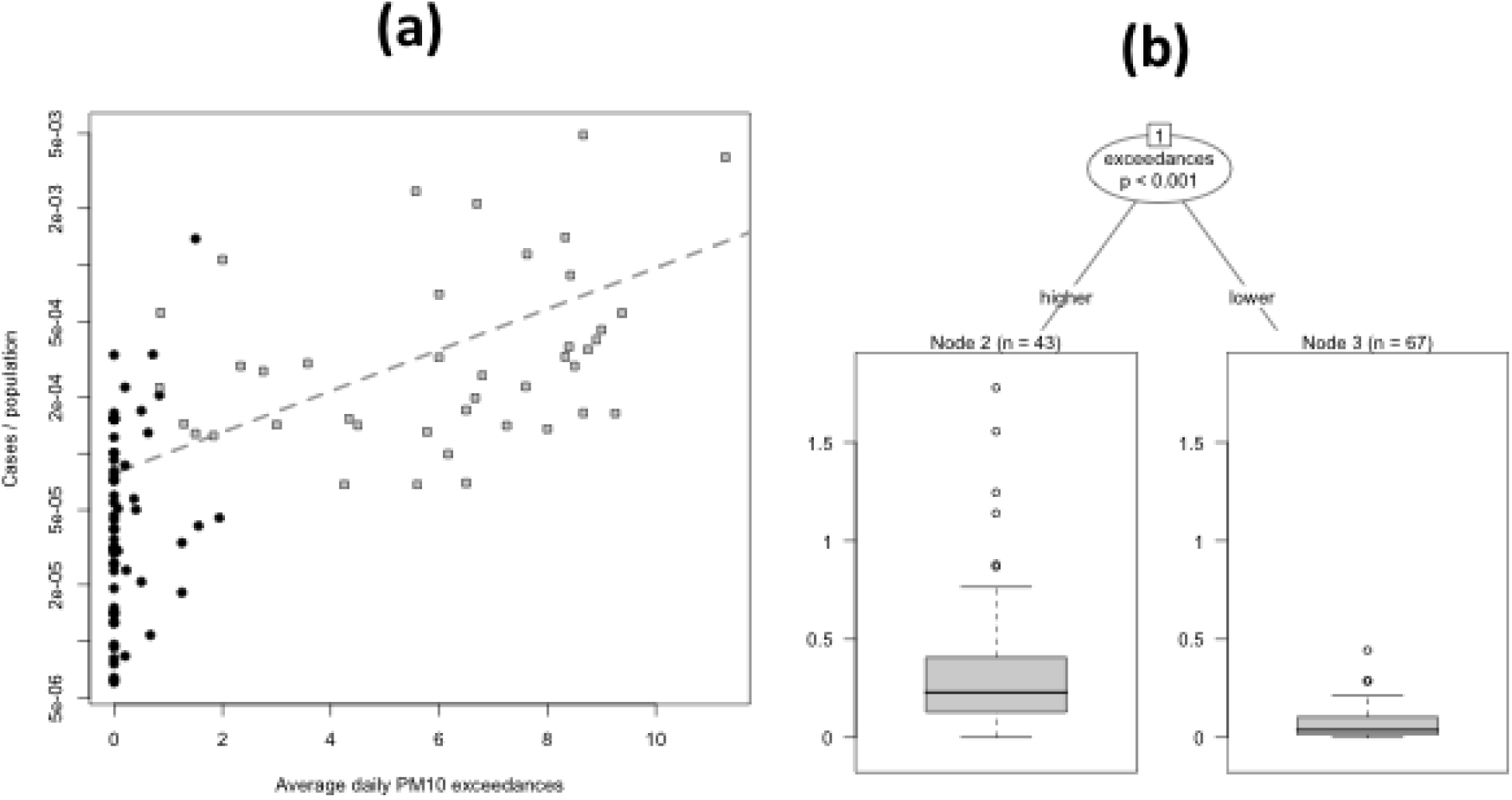
Relationship between the PM_10_ daily limit value exceedances and the COVID-19 cases ratios over Italian Provinces population. **(a)** Scatterplot on semi-logarithmic scale relating the proportion of COVID-19 cases of Northern (gray squares) and Southern (black bullets) Italian Provinces population versus the average of PM_10_ daily limit value exceedances. The dashed binomial (logistic) regression is characterized by an increasing slope of 0.25 (p < 0.001). **(b)** Boxplots showing that - with a 1.29 cut-off value of exceedance - the proportion of COVID-19 cases is greater (p < .001) in most polluted Provinces (39 out of 41 located in Northern Italy) than less polluted Provinces, mainly located in Southern Italy (62 out of 66).

Dividing the Italian peninsula into two areas, the Northern and Southern part along the Tuscan-Emilian Apennines watershed, the exceedances results as follows: 39 of the 41 Northern Provinces falls in the higher PM_10_ category, while on the Southern Provinces the ratio is reversed: 62 over 66 have lower PM_10_ (odds ratio .00, Fisher exact test p < .001).

Also the proportion of commuters over the Province population has a significant (p = 0.01, not depicted) role in predicting the infection rates according to the univariate binary partitioning tree analysis: after setting a cut-off of 47% people daily moving in Provinces, in the Provinces with a lower number of commuters (n = 51) the median infection case over 1000 Province residents is 0.03 (first – third quartile 0.01 – 0.05, range 0.00 – 0.33), while in the other Provinces the median infection cases over 1000 residents is 0.18 (first – third quartile 0.13 – 0.36, range 0.00 – 4.92).

Notably, when performing a bivariate conditional regression exploratory analysis joining both the pollution and the proportion of commuters as possible predictors of the infection rates, one obtains exactly the same tree depicted in Fig. 3b: the commuters proportion loose its effect, suggesting a strong correlation of air quality to the COVID-19 cases percentages breakout.

The logistic regression depicted in Fig. 3a (semi-logarithmic scales) confirms the exploratory analysis: a binomial distributed generalized linear model, corrected for overdispersion, reveals an increasing slope of In order to observe the effect of the particulate matter in big cities having quite similar urbanization, life style, population and number of commuters, Milan and Rome were chosen, finding out that the presence of the first infected people was similar on February 25^th^: 8 and 3 infected persons in Milan and Rome, respectively. However, we considered as the first day of spread for both cities when in Rome the infected persons was about 6 on March 1st (Figure 4).

**Figure 4.**
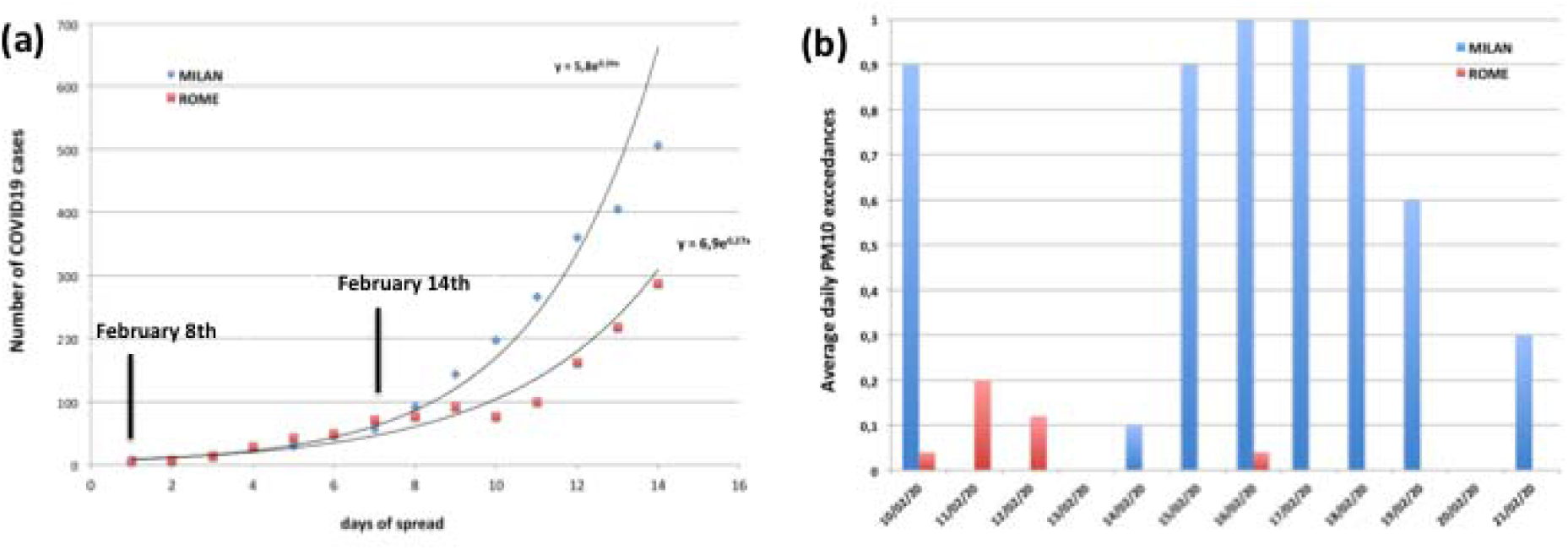
**(a)** Trends of spread in Milan and Rome in the first 14 days of infection; the starting date in Milan is February 25^th^ and could correspond to infections acquired by February 8^th^ that become clinically evident or detectable within 17 days (interval between the infection and diagnosis); Distribution of the average daily PM_10_ exceedances in Rome and Milan on February 2020.

The comparison of the COVID-19 spreads between Milan and Rome showed a higher exponential phase for the former than the latter. However, the trends presented a similar behavior up to 8 days; after the 9 days, the increase of the COVID-19 incidences showed a sudden acceleration of the viral infectivity in Milan. Besides the transmission of SARS-CoV-2 occurring via a close contact with infected people hrough the direct inhalation of liquid droplets emitted by coughs or exhalations and/or by the contact with surfaces contaminated by the virus, the dynamic of COVID-19 incidence observed in Milan with – if compared to that of Rome – suggested also to consider possible route of transmission by airborne route at longer distance. Considering 17 days as average of lag phase, the first day of the infection in Milan that we monitored in February 25^th^ should be referred at the real contamination in February 8^th^. According to this, the acceleration of the COVID-19 incidences in Milan started close to February 14^th^ (Figure 4a) in correspondence to the presence of a large peak of PM_10_ exceedances (Figure 4b) that in Rome was not observed because the start of the incidences was closed to February 13^th^ in a period with the absence of PM exceedances.

The incidence growth rate in Italy was 0,19 per day with a doubling time close to 3.6 days in according with Sanche et al. (2020) who showed a growth rate of infection of COVID-19 in Wuhan, Hubei Province (China) on January 2020 close to 0.21-0.30 per day with a doubling time of 2.3-3.3 [27]. The basic reproductive number (R_0_), estimated by the researchers, was 5.7 consistently with a “super-spread event” by an airborne droplet transmission as described by Wellings and Teunis (2004) for the epidemic curves for Sever Acute Respiratory Sindrome (SARS) during the outbreak on February-June 2003 in Hong Kong, Vietnam, Singapore and Canada [28]. In Rome the growth rate before the lockdown measures (March 13^th^) was 0,27 per day with a doubling time of 2.6 days that were comparable with a “super-spread event” as described for SARS. In Milan the growth rate was significantly higher, close to 0.34 per day with a doubling time of 2.0 days, and suggests a R_0_ value higher than 6.0 quite similar to the epidemic transmission by airborne droplets observed for measles (known to be around 12-18) [29] and to the highest R_0_ estimates documented for China, ranging from 1.4 to 6.49 with a mean of 3.28 and a median of 2.79 (Wuhan: 2.55-2.68; Hubei Province: 6.49; China: 2.2-6.47) [30].

## Discussion

Based on the available literature [2-19], there is enough evidence to consider the airborne route, ant specifically the role of particulate matter, as a possible additional infection “boosting” factor for interpreting the anomalous COVID-19 outbreaks observed in the Northern Italy – known to be one of the European areas characterized by the highest PM concentration [1]. Airborne transmission is certainly more effective in indoor environments, with little ventilation, but it must be considered that the Po Valley, by its atmospheric stability, closely resembles a confined environment and that long-distance virus transport is favored by high concentration of dusts. However, the highly diluted nature of viral bioaerosol in ambient air has been considered a major impediment to viral aerobiological detection –including the investigation of viral interactions with other airborne particles – despite bioaerosol is a well-known factor for the virus transmission via airborne. Recently, Groulx et al. (2018), using an in vitro PM concentrator, suggested that the interaction between airborne viruses and airborne fine particulate matter influence viral stability and infectivity [31]. The stability of aerosol and condensation reactions occur frequently in atmosphere, as organic aerosol change the properties (hygroscopicity, toxicity, optical properties) of other aerosol [32].

Cruz-Sanchez et al. (2013) demonstrated that Respiratory Syncytial virus (RSV) exposed to black carbon, in the form of India ink, prior to co-aerosolization in vitro, and then deposited on a cell substrate, increased viral infectivity [33]. In areas of high vehicle traffic, many different pollutants arising from a variety of sources coexist (car or truck exhausts, emissions from heating installations, etc.) [34], which present a particulate matter emissions containing carbon, ammonium, nitrate and sulfate.

Our findings showed that high frequency of PM_10_ concentration peaks (exceeding 50 µg/m^3^) result in a spread acceleration of COVID-19, suggesting a “boost effect” for the viral infectivity. We found significance differences both in PM_10_ exceedances and COVID-19 spreading between Northern and Southern Italian regions, and we made a focus on Milan and Rome. The infection rate of disease has been higher in Milan, (1.35 million inhabitants, Northern Italy) than in Rome (2.87 million inhabitants, Southern Italy), even if there has not been a substantial difference in urban management and social confinement as well as in ageing index of the two populations. Our research hypothesis is that the acceleration of the growth rate observed in Milan could be attributed to a “boost effect” (a kind of exceptional “super-spread event”) on the viral infectivity of COVID-19, corresponding to the peaks of particulate matter. These first observations suggest that particulate matter could be regarded as an indicator of the severity of COVID-19 infection in terms of diffusion and health outcomes.

The other hypothesis is that PM could act as a carrier for droplet nuclei, triggering a boost effect on the spread of the virus (Figure 5). It could be possible to look at the airborne route of transmission, and specifically to particulate matter, as a “highway” for the viral diffusion, in which the droplet nuclei emitted by the exhalations are stabilized in the air through the coalescence of aerosol with the PM at high concentrations in stability conditions. In fact, the fate of a small droplet of a virus, under normal conditions of clean air and atmospheric turbulence, evaporates and /or disperses quickly in atmosphere. On the contrary in conditions of atmospheric stability and high concentrations of PM, viruses have a high probability of creating clusters with the particles and, by reducing their diffusion coefficient, enhancing their residence time and amount in atmosphere and promoting contagion.

**Figure 5.**
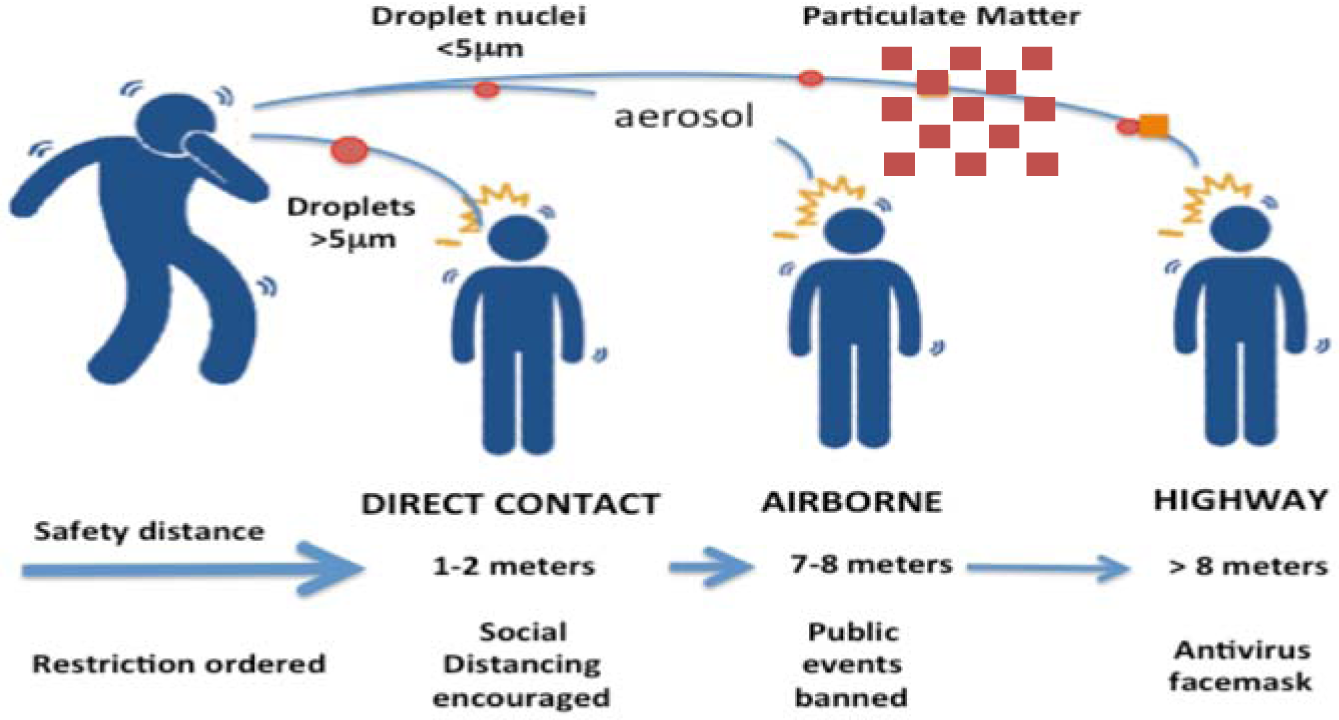
Scheme of possible enhancement of viral transmission through stabilized human exhalation on PM.

Nevertheless, coalescence phenomena require optimal conditions of temperature and humidity to stabilize the aerosols in airborne, around 0-5 °C and 90-100% relative humidity. Recently, Ficetola et al. (2020) showed that the spread of COVID-19 peaked in temperate regions of the Northern Hemisphere with mean temperature of 5°C and humidity of 0.6-1.0 kPa, while decreased in warmer and colder regions [35]. These climatic variables could have a role, together with the presence of high concentrations of particulate matter in the air, in favoring the stabilization of the aerosol in airborne, in line with the model proposed in Fig. 5.

Further experimental studies could confirm the possibility that particulate matter may act as a “carrier” for the viral droplet nuclei, impressing a boost effect for the spreading of the viral infection, as it has been shown for other viruses. Recent studies [36] and recommendation [37] about increased social distancing indicate that a recommended interpersonal distance of significantly more than one meter and usage of personal masks [38] are advisable prevention measures.

It must also be pointed out that long term exposures to high levels of particulate matter itself chronically impair human health and possibly influence clinical course of infections acquired by already debilitated individuals, especially in most vulnerable age groups. Indeed, according to 2005 WHO guidelines, annual average concentrations of PM10 should not exceed 20 μg/m3 (compared to current EU legal limits of 40 μg/m3) and PM2.5 should not exceed 10μg/m3 (compared to current EU legal limits of 25 μg/m3) [39]. Moreover, the exposure-effect relationship between fine particulate matter and health damages is not of linear type, so that it is not really possible to set a threshold below which is foreseeable a complete absence of damage to human health [39].

## Conclusion

The available literature on the role of airborne transmission, and this first preliminary observation of consistent association between the number of COVID-19 infected people and PM_10_ peaks, points out the opportunity of a further computational and experimental research on this route of transmission, and the potential role of PM on viral spread and infectivity (in addition to the possibility of regarding PM levels as an “indicator” of the expected impact of COVID-19 in most polluted areas). There is the rational for carrying out experimental studies specifically aimed at confirming or excluding the presence of the SARS-CoV-2 and its potential virulence on particulate matter of Italian cities as well as at European and international level.

Urgent actions must be adopted to counteract climate changes and the alteration of ecosystems that might trigger new and unexpected threats to human health such as that of COVID-19, which we are so dramatically experiencing worldwide.

## Data Availability

Data will be made available upon reasonable requests

## Authors Contributions

L. Setti, F. Passarini, G. De Gennaro^3^ P. Barbieri, M.G. Perrone, A. Piazzalunga, M. Borelli, J. Palmisani, A. Di Gilio, P. Piscitelli, A. Miani conceived, prepared, wrote and revised the manuscript.

